# Seroprevalence of IgG antibody against SARS-CoV-2 among health care workers of anaesthesia departments from various hospital settings in India

**DOI:** 10.1101/2021.03.20.21253819

**Authors:** Debaprasad Parai, Hari Ram Choudhary, Girish Chandra Dash, Annalisha Peter, Dipika Saket, Ritesh Roy, Gaurav Agarwal, Saroj Sahoo, Usha Kiran Rout, Rashmi Ranjan Nanda, Sanghamitra Pati, Debdutta Bhattacharya

## Abstract

**Background:** Health care workers (HCWs) are the most susceptible group to get COVID-19 infection and this group always need special attention as they are the key human resource to contain this pandemic.

**Objective:** To track down the seroprevalence among a particular group of HCWs working in the anaesthesia department in hospital settings.

**Study design:** Two rounds of serosurvey were done to track the dynamicity among the 128 and 164 HCWs participants in the first round and second round, respectively. 5 mL of blood was collected and IgG SARS-CoV-2 antibody was tested in Abbott Architect i1000SR.

**Results:** The seroprevalence found in the first and second round was 12.5% and 38.4%, respectively. A significant number (n=61, 77.21%) of seropositivity came from the asymptomatic HCWs group as found in both the survey. There was no significant association among different age, gender and RT-PCR tested groups.

**Conclusion:** Routine diagnosis of COVID-19 should be referred among HCWs to identify and act upon unrecognized SARS-CoV-2 infection.

## Introduction

The first incidence of coronavirus disease-19 (COVID-19) was found in December 2019 from Wuhan city of China (1). Since then, the cumulative new cases are being reported from each part of the world which forced the world health organization (WHO) to declare the disease as a pandemic in March 2020 (2). A total number of 81.5 million confirmed cases has been registered and the global death count reached 1.8 million by the end of 2020. India, the second largest populated country has seen a positive case above 10 million and mortality of 100,000 in a single year (3). The clinical manifestation of severe acute respiratory syndrome coronavirus 2 (SARS-CoV-2) infection mainly varied from very mild to severe pneumonia, respiratory distress and death. However, as most of the patients develop no symptoms, the exact number of positive cases should be far more than the registered data (4).

Health care workers (HCWs) from anaesthesia departments of a hospital are one of the most vulnerable groups in the transmission of COVID-19 through patients, colleagues or from the community. Although real time polymerase chain reaction (RT-PCR) of nasopharyngeal swab sample is the current gold standard for the confirmation of SARS-CoV-2 infection, screening of symptoms for HWCs and then go for RT-PCR is a standard practice to control infection (5, 6). However, the high proportion of asymptomatic or pre-symptomatic cases often neglect the potential disease transmitter which aggravates the infection among patients and other HCWs (7). The current study was to determine the utility of a regular serological survey for the screening of asymptomatic infected individual among HCWs related to patient’s anaesthesia in hospital settings.

## Methods

### Study settings

A total of 128 HCWs participated in the first serological survey in September 2020 and 164 HCWs in the second survey conducted in December 2020 from 12 hospitals in Bhubaneswar, India. All the participants were working in the anaesthesia department and responsible to deliver care and services to critical patients, either directly as physicians or nurses, or indirectly as technicians, administrative staffs or other support staffs. Only the willing participants were included in this study and written informed consent was obtained from each participant. Clinical and other demographic data was collected during sample collection. The study protocol was approved by the Institutional Human Ethics Committee of the Indian Council of Medical Research-Regional Medical Research Centre, Bhubaneswar.

### Collection of blood

Sera samples were collected and tested to determine the presence of IgG antibodies against the nucleocapsid protein of SARS-CoV-2 in an automated analyzer ARCHITECT i1000SR (Abbott Laboratories, Chicago, USA) using chemiluminescent microparticle immunoassay (CMIA) technology. The assay specificity in this platform was 99.63% (95% confidence of interval [CI]: 99.05-99.90%) and sensitivity was 100% (95% CI: 95.89-100%) when tested after 14 days of symptom onset as per the manufacturer. The cut-off value was 1.4 index.

### Statistical analysis

Descriptive statistical analyses were performed by SPSS software (IBM SPSS Statistics for Windows, version 24.0, Armonk, NY) and GraphPad Prism software version 7.0 for Windows (GraphPad Software, La Jolla, California, USA). *p* values < 0.05 was considered as significant.

## Results

In the first round of the survey, 16 HCWs were found to have antibody which is only 12.5% of the total participants. The seropositive number increased to 63 from a total of 164 HCWs in the second serosurvey which is around 38.4% (Figure 1). We analyzed and correlated the antibody titre values from all the 79 seropositive individuals against symptoms status, age groups and gender (Table 1). A total of 35 HCWs (11.98%) developed symptoms out of 292 participants throughout two survey phases with three HCWs requiring hospitalization. Among antibody-positive HCWs, only 18 persons (22.78%) developed any COVID symptoms within 30 days of the survey which is statistically significant with the asymptomatic antibody-positive group (77.22%). The mostly found symptoms among this group were fever (83.33%) followed by cough (50%), malaise (27.7%), fatigue (27.7%), throat pain (22.2%), loss of smell (22.2%), chest pain (16.7%). Nasopharyngeal or oropharyngeal samples from 90.63% (first survey) and 92.07% (second survey) of the HCWs had been tested in RT-PCR but only 22 (8.24%) were tested positive in RT-PCR. In both the serosurvey, higher seroprevalence was found in the male group compared to the female although it was statistically non-significant. In this study, we categorised four different groups based on patients’ age *viz*. group I (≤29 years), group II (30-45 years), group III (46-60 years) and group IV (>60 years). Contingency chi-square test analysis couldn’t find any significant correlation among the different age groups throughout both the survey. The highest percentage of seropositivity was found in the 45-59 years age group in the first round (27.3%) whereas, in the second round, the maximum seropositive came from 30-44 years (43.0%). In the first round of the survey, 116 HCWs were already tested in RT-PCR and the second phase, the RT-PCR tested number was 151. A total of 5 and 17 HCWs were confirmed RT-PCR positive in the first and second serosurvey, respectively. Among those 22 cases, 19 individuals (86.36%) developed antibody as found in this study (Table 1).

**Table 1.** Demographic characteristics, testing status and symptom profile of the study participants and the distribution of seroprevalence.

| Variables |  | Round 1 |  |  | Round 2 |  |  |
| --- | --- | --- | --- | --- | --- | --- | --- |
|  |  | N= | Positive= (% , 95% CI) | Chi Square (p-Value) | N= | Positive= (% , 95% CI) | Chi Square (p-Value) |
| **Age Groups** | **≤ 29** | 34 | 4<br>(11.8 % , 3.3-27.5%) | 3.115<br>(0.374) | 63 | 22<br>(34.9 % , 23.3-48%) | 2.244<br>(0.523) |
|  | **30-44** | 77 | 9<br>(11.7 % , 5.5-21%) |  | 79 | 34<br>(43 % , 31.9-54.7%) |  |
|  | **45- 59** | 11 | 3<br>(27.3 % , 6-61%) |  | 16 | 6<br>(37.5 % , 15.2-64.6%) |  |
|  | **≥ 60** | 6 | 0<br>(0 % , 0-0%) |  | 6 | 1<br>(16.7 % , 0.4-64.1%) |  |
| **Sex** | **Male** | 76 | 10<br>(13.2 % , 6.5-22.9%) | 0.074<br>(0.786) | 91 | 39<br>(42.9 % , 32.5-53.7%) | 1.705<br>(0.192) |
|  | **Female** | 52 | 6<br>(11.5 % , 0.4-23.4%) |  | 73 | 24<br>(32.9 % , 22.3-44.9%) |  |
| **Symptomatic** | **Yes** | 13 | 4<br>(30.8 % , 9.1-61.4%) | 4.415<br>(0.036) | 22 | 14<br>(63.6 % , 40.7-82.8%) | 6.832<br>(0.009) |
|  | **No** | 115 | 12<br>(10.4 % , 5.5-17.5%) |  | 142 | 49<br>(34.5 % , 26.7-42.9%) |  |
| **RT PCR Positive** | **Yes** | 5 | 4<br>(80 % , 28.4-99.5%) | 20.876<br>(0.000) | 17 | 15<br>(88.2 % , 63.6-98.5%) | 18.207<br>(0.000) |
|  | **No** | 111 | 11<br>(9.9 % , 5.1-17%) |  | 134 | 46 (34.3 % , 26.3-43%) |  |
| **RT-PCR Tested** | **Yes** | 116 | 15<br>(12.9 % , 7.4-20.4%) | 0.210<br>(0.647) | 151 | 61<br>(40.4 % , 32.5-48.7%) | 3.165<br>(0.075) |
|  | **No** | 12 | 1<br>(8.3 % , 0.2-38.5%) |  | 13 | 2<br>(15.4 % , 1.9-45.4%) |  |
| **Total** |  | 128 | 16<br>(12.5 % , 7.3-19.5%) | - | 164 | 63<br>(38.4 % , 30.9-46.3%) | - |

**Figure 1.**
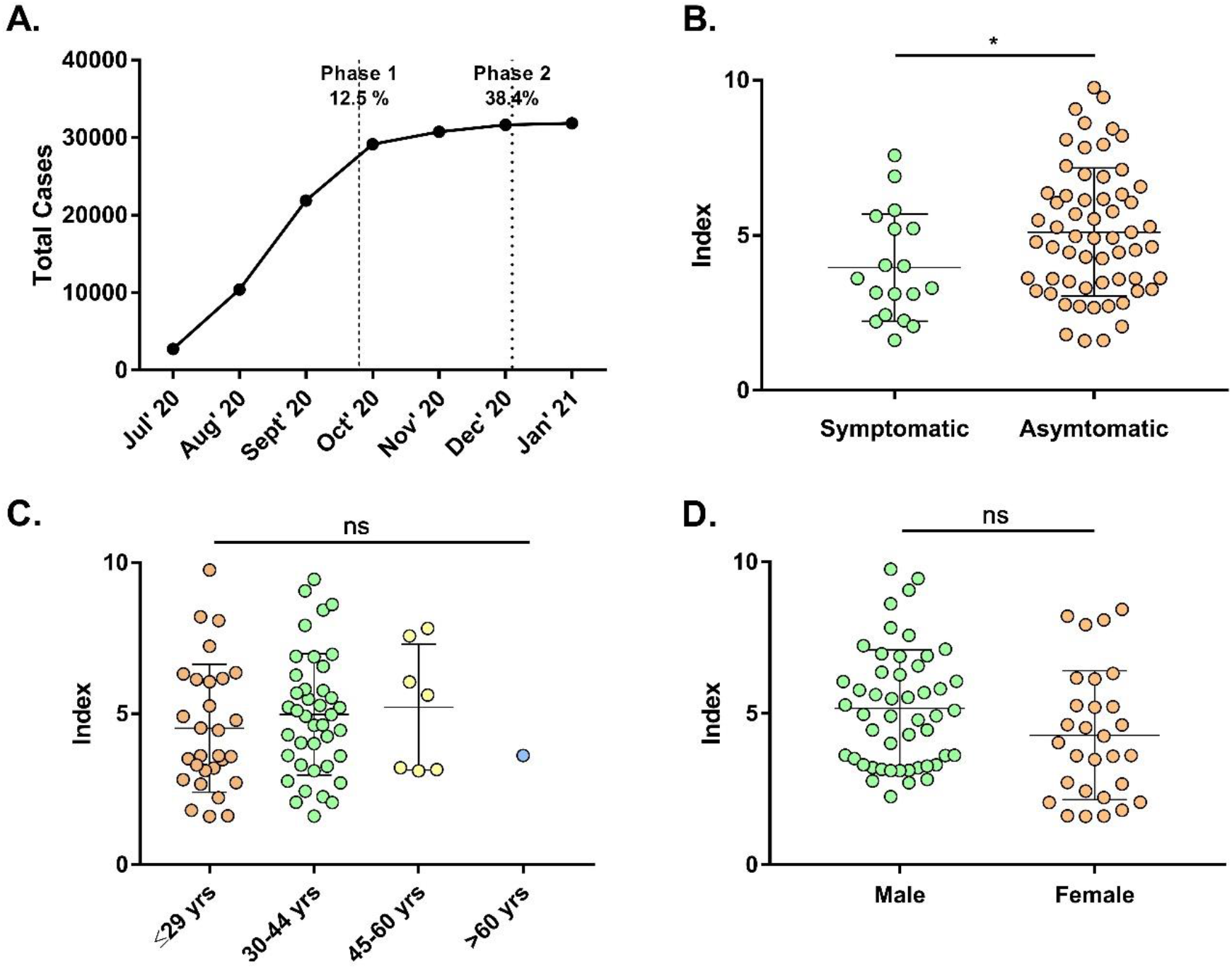
Seroprevalence and their relationship between different categorical variables. (A) Average of total monthly cases and point of serosurvey with seroprevalence. Antibody titre and its relationship with symptom status (B), age groups (C) and gender (D). p <0.05 was considered as statistically significant.

## Discussion

Early diagnosis and understanding of immunological responses are essential to study the COVID-19 disease progression. Serological studies provide relevant information about the spread of infection among the communities and the number of individuals who have infected in the recent past. Conducting such studies among the health care workers regardless of the onset of the symptoms is important to understand the level of exposure within the hospitals and this helps us to identify and prevent the spread of infection. Similarly, the information about the recently infected HCWs could be helpful for the hospital administration to skip the compulsory quarantine period and can be useful for maximizing the use of an available resource or manpower (5, 8).

Seroprevalence found in both the survey among HCWs was higher than the 2^nd^ and 3^rd^ national serosurvey conducted in India during August to September 2020 and December 2020 to January 2021 where the seropositivity found among the adults were 6.6% and 21.4%, respectively (9). The total number of confirmed COVID-19 cases in Bhubaneswar was 10,425 at the end of August 2020 but it reached a peak of 30,751 by the end of November 2020 (10). The rapid increase in the total confirmed cases can be easily correlated with the difference in the seroprevalence between the two serosurveys among the HCWs. Our study also found that most of the seropositive HCWs were asymptomatic and the ratio was 3.4:1 to the asymptomatic cases. Moreover, most of the HCWs (n=277) were tested in RT-PCR but only 7.94% (n=22) were confirmed as positive. Seropositive significance in chi-square test among symptomatic and asymptomatic suggests a need for routine screening of COVID-19 is mostly needed as this group is the most vulnerable one in a hospital setting. This study from the anaesthesia department HCWs can easily be correlated with some other findings on overall hospital workers (11, 12).

Our study also has a few limitations. First, as our participants were from the anaesthesia department of hospital settings, thus it may not speculate the exact association of immune response among the whole HCW community. Second, underestimation during reporting of the symptoms might be happened due to the immense workload of health workers which force them to neglect mild symptoms and timely diagnosis.

In conclusion, health workers are the most susceptible group to get COVID infection. Conditions like asymptomatic or pre-symptomatic make the scenario complicated due to the non-occurrence of routine diagnosis. Although our study has few limitations, there is no such study with health workers only from the anaesthesia department of hospital settings to demonstrate the seroprevalence and its association with symptoms, RT-PCR test, age and gender to our best knowledge. Our result concludes that HCWs should routinely get tested for COVID-19 as most of them may be asymptomatic or pre-symptomatic which indeed a much more concerning situation when it comes to patients’ safety.

## Data Availability

The data is available with the corresponding author and can be provided on valid request.

## Ethics approval and consent to participate

The study was approved by the Institutional Human Ethics Committee of ICMR – RMRC Bhubaneswar.

## Acknowledgement

The authors gratefully acknowledge all the healthcare workers from the Indian Society of Anaesthesiology, Bhubaneswar Branch for their tireless dedication at each level to fight COVID-19. The authors are thankful to the Indian Council of Medical Research, New Delhi and Dept. of Health & Family Welfare, Govt. of Odisha, for providing financial support for this study.

## References

1. C. Wang, P.W. Horby, F.G. Hayden, G.F. Gao, A novel coronavirus outbreak of global health concern, The Lancet. 395 (2020) 470–473.

2. World Health Organization. WHO Director-General’s opening remarks at the media briefing on COVID-19. https://www.who.int/director-general/speeches/detail/whodirector-general-s-opening-remarks-at-the-media-briefing-on-covid-1911-march2020

3. COVID Dashboard. https://www.mohfw.gov.in/

4. R. Li, S. Pei, B. Chen, Y. Song, T. Zhang, W. Yang, et al, Substantial undocumented infection facilitates the rapid dissemination of novel coronavirus (SARS-CoV-2), Science. 368 (2020) 489–93.

5. J.R.M. Black, C. Bailey, J. Przewrocka, K.K. Dijkstra, C. Swanton, COVID-19: the case for health-care worker screening to prevent hospital transmission, Lancet. 395 (2020) 1418–20.

6. V.M. Corman, O. Landt, M. Kaiser, R. Molenkamp, A. Meijer, D.K. Chu, et al, Detection of 2019 novel coronavirus (2019-nCoV) by real-time RT-PCR, Euro. Surveill. 25 (2020) 2000045.

7. V.H. Ferreira, A. Chruscinski, V. Kulasingam, T.J. Pugh, T. Dus, B. Wouters, et al, Prospective observational study and serosurvey of SARS-CoV-2 infection in asymptomatic healthcare workers at a Canadian tertiary care center, PLoS ONE. 16 (2020) e0247258.

8. L. Fernández-Barat, R. López-Aladid, A. Torres, The value of serology testing to manage SARS-CoV-2 infections, Eur. Respir. J. 56 (2020) 2002411.

9. M. Banaji. COVID-19: What the Third National Sero-Survey Result Does and Doesn’t Tell Us. https://science.thewire.in/health/third-national-seroprevalence-survey-icmr-covid-19-rural-prevalence-test-positivity/, 2021

10. -COVID Dashboard. https://statedashboard.odisha.gov.in/

11. A.L. Garcia-Basteiro, G. Moncunill, M. Tortajada, M. Vidal, C. Guinovart, A. Jiménez, et al, Seroprevalence of antibodies against SARS-CoV-2 among health care workers in a large Spanish reference hospital, Nat. Commun. 11(2020) 3500.

12. A. Kantele, T. Lääveri, L. Kareinen, S.H. Pakkanen, K. Blomgren, S. Mero, et al, SARS-CoV-2 infections among healthcare workers at Helsinki University Hospital, Finland, spring 2020: Serosurvey, symptoms and risk factors, Travel Med. Infect. Dis. 39 (2021) 101949.

